# The Cost-effectiveness of One-step Point-of-care Hepatitis C Virus RNA Testing in the United States

**DOI:** 10.1101/2025.09.27.25336816

**Authors:** Neda Al Rawashdh, Cheryl P Ferrufino, Mindy M Cheng, Shivang Doshi, Emily B Kahn

## Abstract

**Background:** The standard of care (SOC) two-step laboratory-based hepatitis C virus (HCV) diagnostic algorithm is associated with high rates of undiagnosed cases, patients lost to follow-up, and low rates of treatment initiation. This study examines the cost-effectiveness of a one-step point-of-care HCV RNA diagnostic strategy (POC_RNA) compared to SOC across distinct settings of care serving people at high-risk of HCV infection.

**Methods:** A hybrid decision tree and HCV transmission model was developed to estimate the clinical and economic outcomes of HCV testing strategies in community health centers (CHC), emergency departments (ED), harm reduction clinics (HRC), and mobile outreach / street medicine programs (MO/SM). The model analyzed costs from a U.S. third-party payer perspective and projected outcomes over a 1-year and lifetime horizons. Model inputs were sourced from real-world claims data and the published literature. One-way and probabilistic sensitivity analyses were performed to assess the impact of uncertainty on results.

**Results:** In the base-case analysis, one-step POC_RNA was projected to increase rates of diagnosis, linkage to care, treatment initiation, successful treatment, and reduce first-generation transmitted cases when compared to SOC across all settings. Although one-step POC_RNA was estimated to increase initial treatment costs in the CRC, HRC, and MO/SM settings, these costs were offset in the longer-term by average savings from reduced incidence of chronic HCV infections and complications.

**Discussion:** This study suggests that a one-step POC HCV RNA testing strategy is optimal for high-risk populations and a critical component of HCV elimination efforts.

## Background

Hepatitis C virus (HCV), one of the most common types of viral hepatitis, is an infection that primarily affects the liver. If left untreated, HCV infection can lead to liver fibrosis, cirrhosis, and liver cancer. HCV remains a leading cause of liver transplants in the United States (U.S.). It has been estimated that between 2.4 – 4.7 million people in the U.S. have active HCV infection; approximately 75-85% of cases are asymptomatic and more than 50% of cases are unaware of being infected.^1, 2^ Notably, the estimated annual number of acute HCV infections has increased 492% between 2010 to 2021, with the majority of new HCV infections associated with injection drug use, especially among young adults.^3^ Despite the availability of highly effective oral direct-acting antiviral (DAA) agents introduced in 2013, the incidence of HCV infection continues to rise.^2^ Effective HCV treatments alone are insufficient to reduce the incidence of HCV in the absence of a robust and simplified screening and diagnostic paradigm.^4, 5^

The standard of care (SOC) for diagnosing HCV infection is a two-step laboratory-based diagnostic testing algorithm in which the first step is a HCV antibody test, followed by a confirmatory HCV RNA quantitative test for persons with detectable HCV antibodies.^6^ The two-step SOC algorithm typically requires more than a single clinic and/or lab visit for an infected person to be diagnosed and subsequently linked to care and initiated on treatment.

Linkage to care (LTC) is predicated on successful contact and call-back of persons who have a positive HCV RNA test and thus increases the risk of patients becoming lost to follow-up (LTFU), especially among high-risk populations such as people who inject drugs (PWID). LTC rates have been reported ranging from 36-65% among PWID. Consequently, the percentage of people who are diagnosed with HCV and initiate treatment remains low due to high rates of LTFU.^5^

Individuals who were recently infected and have not yet seroconverted are at risk of being missed with the two-step SOC algorithm because it typically takes 8 to 11 weeks (and up to 6 months) to mount a detectable antibody response.^6^ People in this early phase of infection may be unaware of their infection and can transmit the virus to others.

Challenges with completion of a two-step diagnostic algorithm and subsequent treatment are exacerbated by the reality that many high-risk populations do not present at traditional healthcare settings or engage in routine healthcare.^6^ To address challenges that lead to LTFU, the American Association for the Study of Liver Diseases (AASLD) and the Infectious Diseases Society of America (IDSA) recently released the Point of Care (POC) Test and Treat Algorithm (https://www.hcvguidelines.org/), which recommends the use of POC HCV RNA testing at the initial visit as well as a simplified assessment for treatment initiation. An “RNA first” test strategy simplifies diagnosis of HCV infection and enables faster LTC and treatment, potentially in a single encounter. Faster LTC is expected to reduce LTFU and increase the rate of treatment initiation among those diagnosed with HCV.

In June 2024, the U.S. Food and Drug Administration granted marketing authorization for the first POC HCV RNA test in the U.S. The Xpert^®^ HCV test (Cepheid, Sunnyvale, CA) is an automated *in vitro* reverse transcription polymerase chain reaction (Rt-PCR) test for the qualitative detection of HCV RNA in capillary whole blood from a fingerstick. The Xpert^®^ HCV test provides results in one hour or less and may be performed in healthcare settings operating under a Clinical Laboratory Improvement Amendments (CLIA) Certificate of Waiver, enabling testing in non-traditional care settings and facilitating rapid HCV diagnosis without the need for phlebotomy.^7^

The objective of this analysis is to evaluate the costs and outcomes of HCV RNA-first testing at the POC as part of a one-step test-and-treat strategy (POC_RNA) across various care settings compared to the SOC.

## Methods

### Cost-Effectiveness Model Structure

A hybrid decision tree (Figure 1a) and HCV transmission model (Figure 1b) was developed using Excel 365 (Microsoft, Bellevue, WA) to evaluate the clinical and economic outcomes of HCV testing strategies in four distinct settings where high-risk populations, primarily PWID, present for healthcare and/or services (community health centers (CHC), emergency departments (ED), harm reduction centers (HRC) (including opioid treatment programs, syringe service programs, and substance use disorder treatment clinics), and mobile outreach/street medicine (MO/SM) programs). The model analyzed costs from a U.S. third-party payer perspective (weighted average of commercial payer, Medicare, and Medicaid by setting) using a hypothetical cohort of 10,000 people eligible for HCV testing within each setting. A decision tree structure was used to model the patient care pathway of the cohort in each setting from diagnostic testing to treatment, cure, and long-term outcomes, while the HCV transmission model projected viral transmission.

**Figure 1a:**
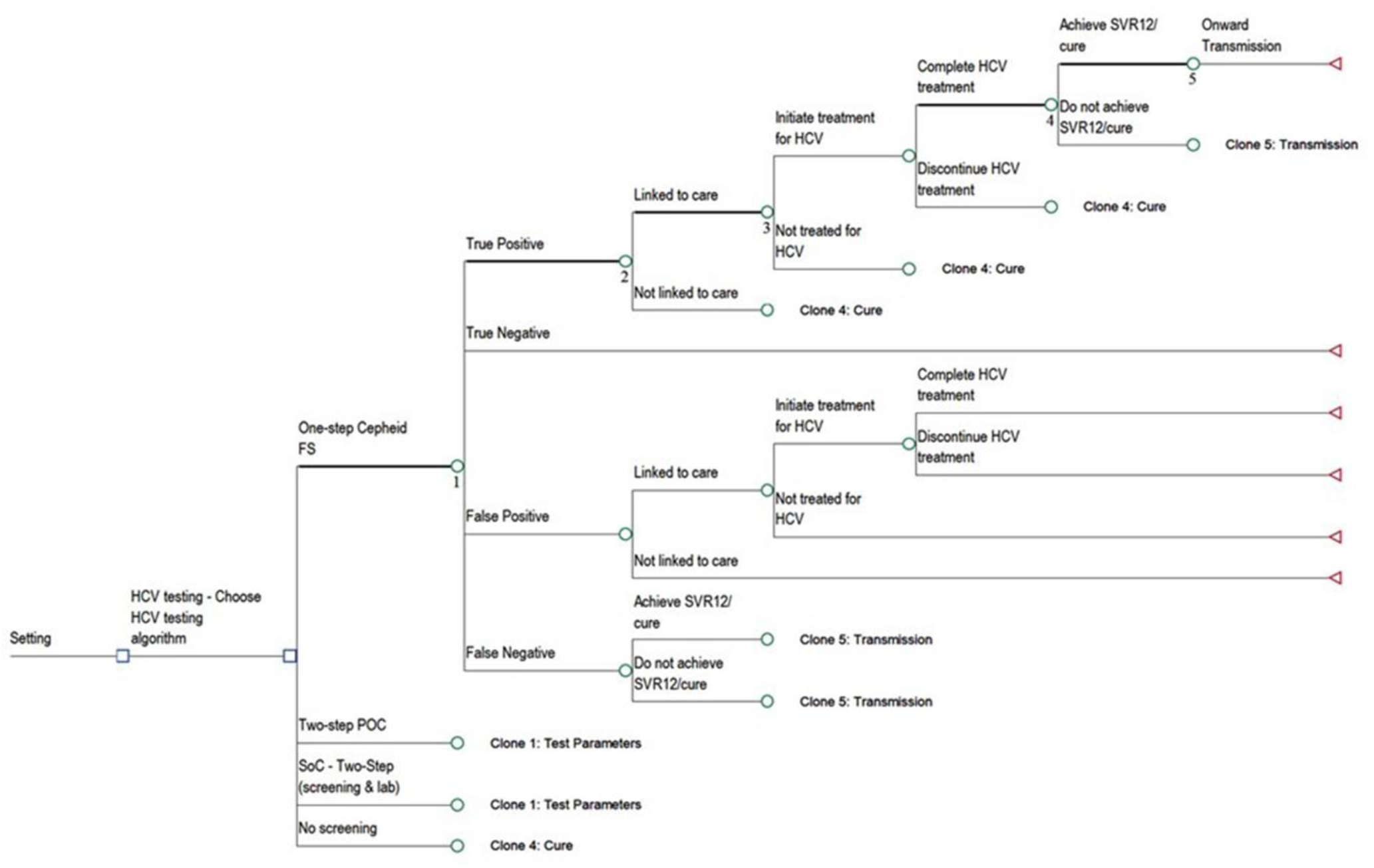
Decision tree model

**Figure 1b:**
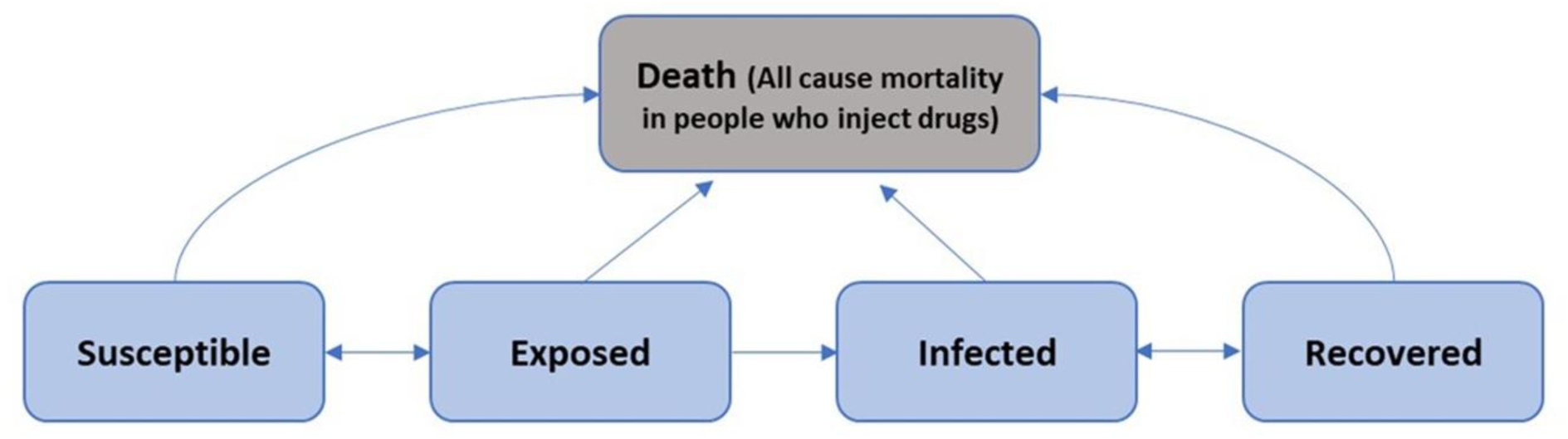
Disease transmission model

The model incorporated a one-year time horizon to capture immediate “test-to-treat” outcomes: HCV infections diagnosed, percentage of cases linked to care, initiated on treatment, and achieved sustained virologic response 12 weeks post-treatment (SVR12), and forward transmission to secondary cases. The model also assessed longer-term impacts of successful “test-and-treat” on averting chronic hepatitis C and long-term complications (cirrhosis, hepatocellular carcinoma (HCC)), death rates, life-years (LYs) and quality-adjusted life years (QALYs) gained. Additionally, total and incremental costs, as well as incremental cost-effectiveness ratios (ICER) for each clinical outcome were calculated.

LTC is defined as patients attending their first follow-up visit after diagnosis, while treatment initiation is marked by the filling of the first prescription for treatment. Initiation of treatment among people diagnosed using POC testing was assumed to be 32% higher than among people tested with SOC, based on estimates derived from the literature.^8^ The model assumes that people receiving either negative HCV antibody or RNA results do not undergo further HCV testing. For those who are diagnosed, but not linked to care or initiated on treatment, a 30% probability of spontaneous recovery was considered, with the remaining cohort at risk of developing chronic hepatitis C and transmitting the virus to others.^9^ Persons with active HCV infections who do not achieve SVR12 are returned to the “susceptible” compartment in the Susceptible-Exposed-Infectious-Recovered disease transmission model, using a reproductive number (R0) of 2.99.^10^

### Model Inputs

#### Clinical inputs

The prevalence of HCV antibodies and HCV RNA among people with and without HCV antibody were obtained from the published literature for each setting of care. The overall prevalence of HCV infection for the SOC was calculated as [*prevalence of HCV antibodies x prevalence of HCV RNA]*. However, for the one-step POC_RNA strategy, the overall HCV infection prevalence was adjusted to include those who tested negative for the HCV antibody but had positive RNA results (i.e., acute infections). The setting-specific prevalence of HCV RNA was used to calculate the number of HCV cases detected with each testing strategy. Test sensitivity was applied to HCV RNA prevalence to calculate the number of true positive and false negative results, while test specificity was used to calculate the number of true negative and false positive results among high-risk individuals without HCV infection. (Table 1)

**Table 1:**
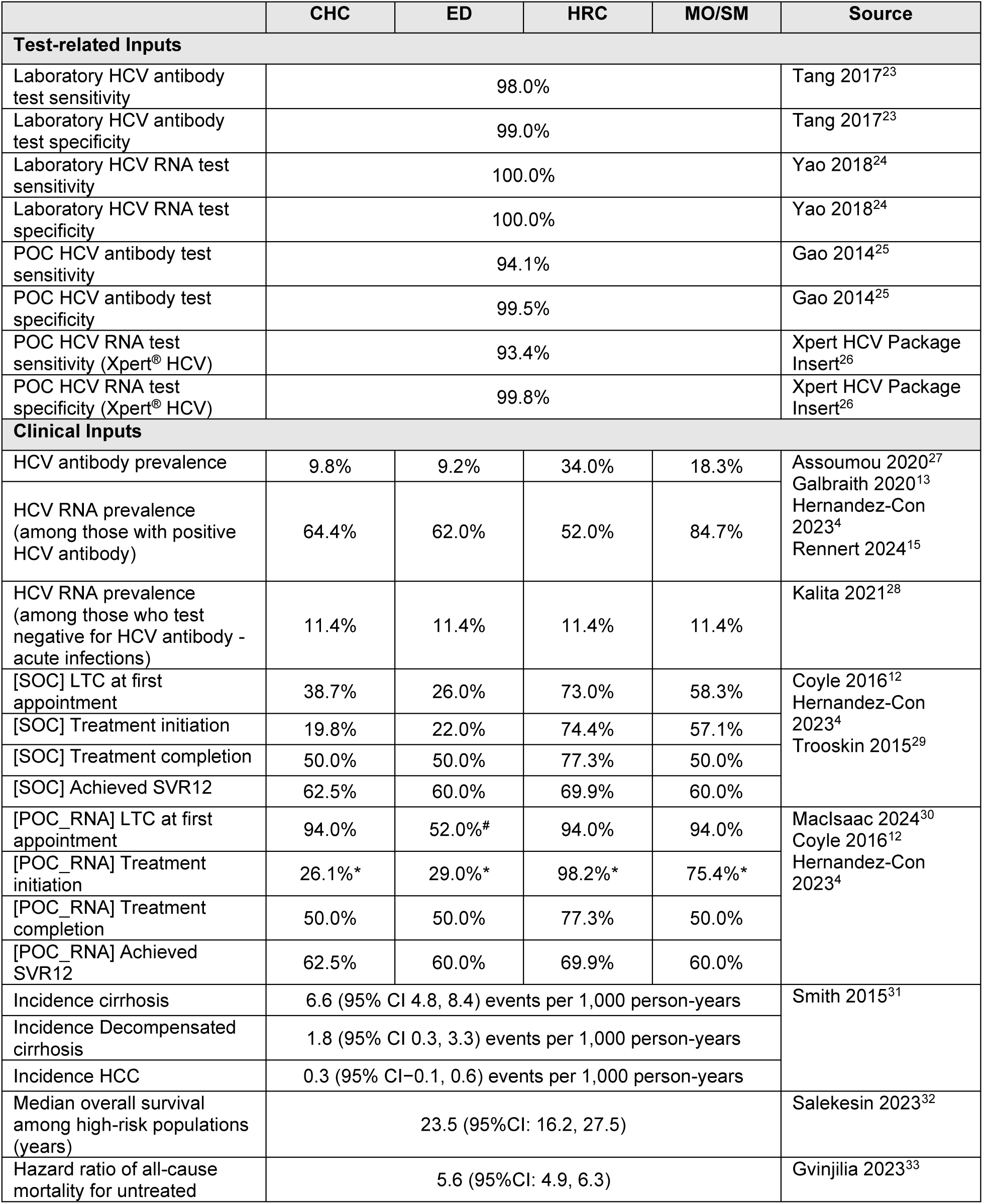
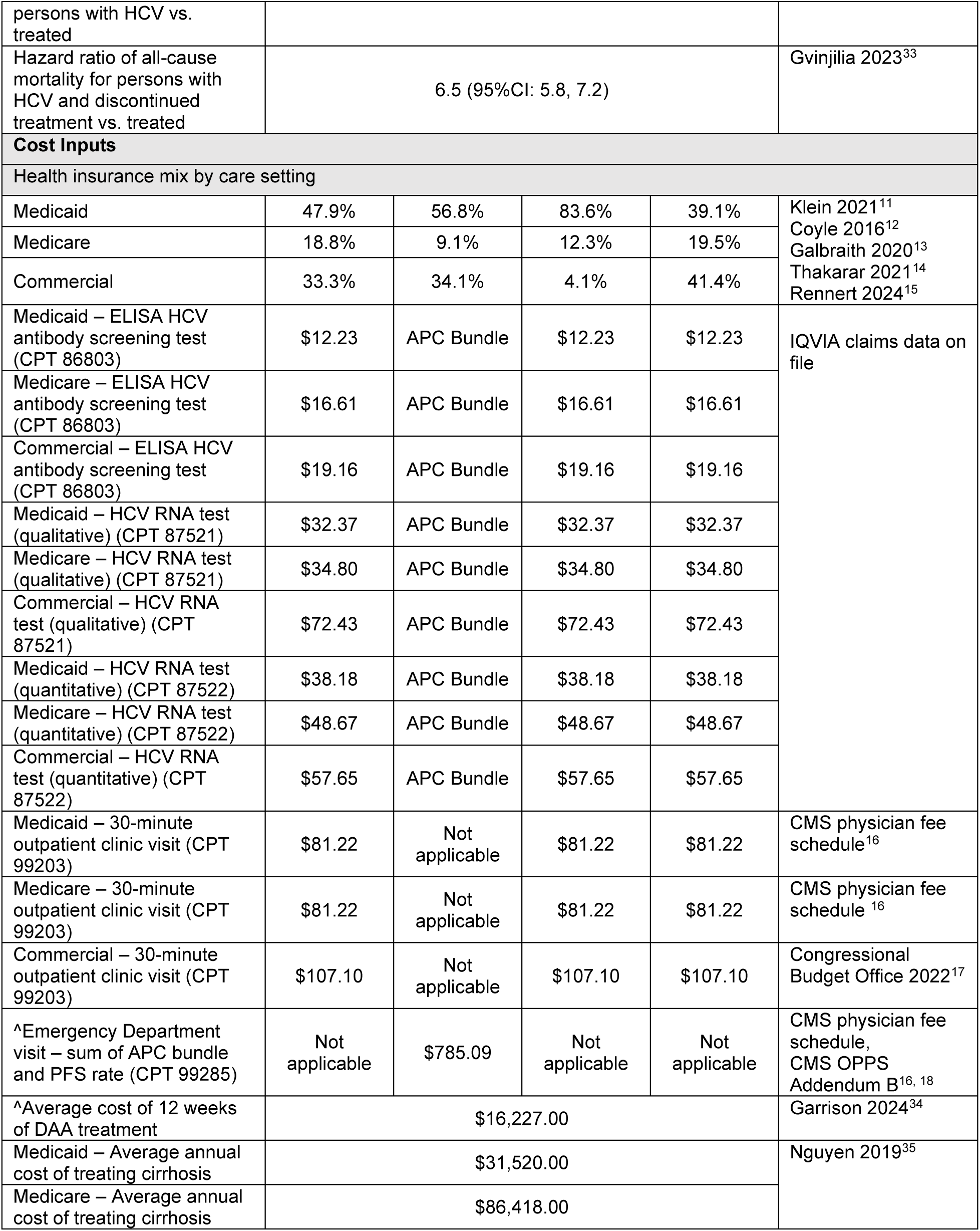
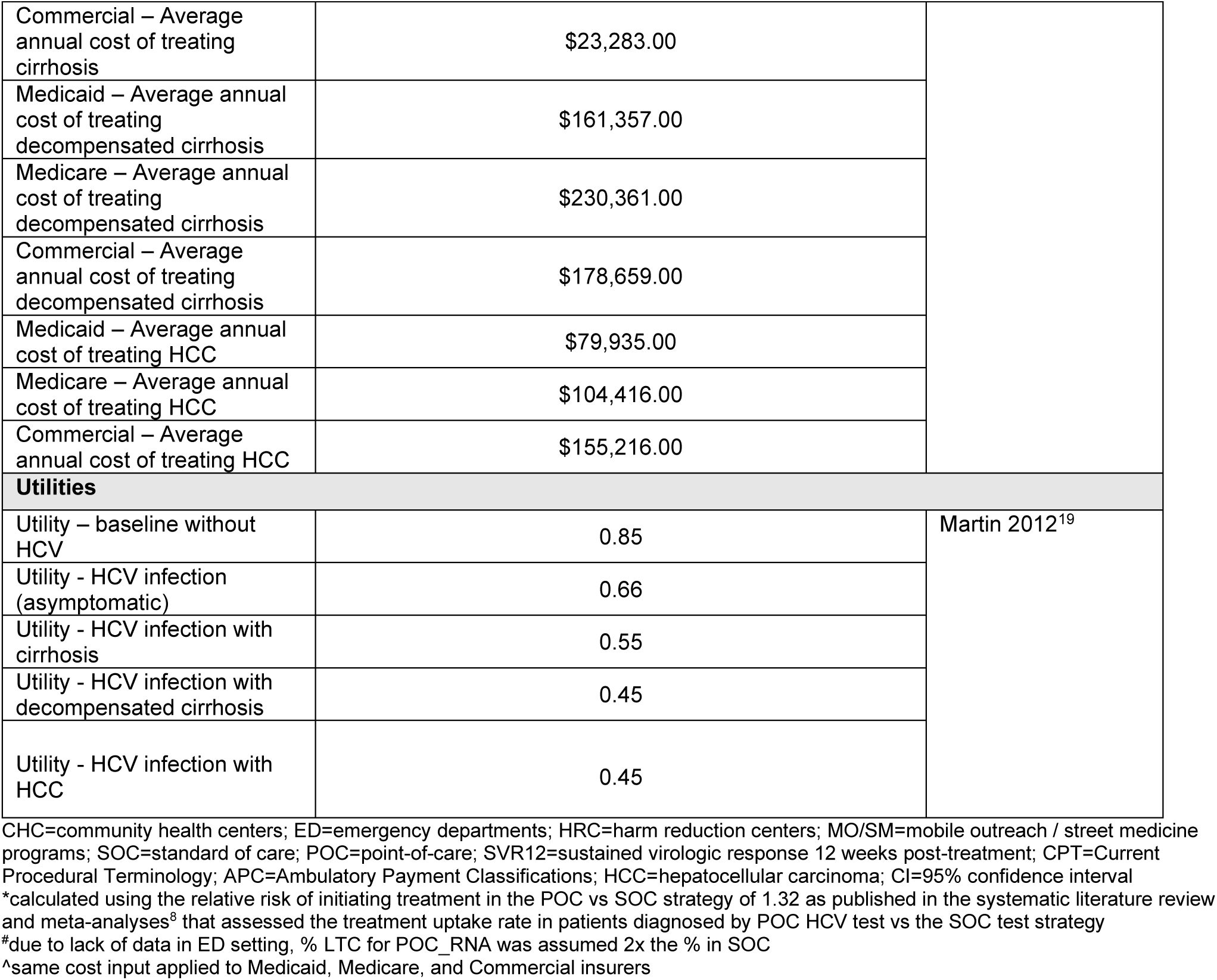
Model Inputs

Persons with positive HCV RNA results, including true positives and false positives who were linked to care, were assumed to receive oral DAA treatment for 12 weeks. The percentages of people with a positive diagnosis at each step of the HCV care cascade (linked to care, initiating treatment, completing treatment, and achieving SVR12) were derived using setting-specific estimates from the literature for the SOC and POC_RNA strategies. Due to lacking data demonstrating implementation of POC_RNA in an ED setting, the percentage of diagnosed patients successfully linked to care in ED was assumed to be double that in the SOC for the base-case analysis and varied +/-20% in sensitivity analysis. (Table 1)

Conversely, those with negative (true or false negative) antibody or RNA results did not enter the HCV care cascade; those with false negative results were at risk of developing long-term complications from chronic infection and transmitting the virus to others. The percentages of people who developed chronic hepatitis C, cirrhosis, and HCC were calculated using published incidence rates among untreated PWID with HCV infection. Survival outcomes, including median survival time among PWID and hazard ratios of death among the untreated population versus the successfully treated population, were fit to exponential distributions to calculate the LYs and QALYs from each testing strategy. (Table 1)

#### Costs and utility inputs

The total cost for each comparator was calculated in each setting (CHC, ED, HRC, MO/SM) based on literature-derived weighted distributions of insurance coverage (Medicare, Medicaid, commercial payer) among the model cohort presenting at each setting.^11–15^ (Table 1) For the one-year time horizon analysis, direct medical costs associated with diagnostic testing and treatment in the CHC, HRC, and MO/SM care settings were based on reimbursement rates for HCV antibody and RNA tests, a 30-minute outpatient visit, and the average cost of 12 weeks of oral DAA treatment. Annual management costs of long-term complications were added for the life-time horizon analysis and assumed care would be provided across different settings, not necessarily in the original setting where HCV testing was performed. Reimbursement rates under a fee for service model were applied to the CHC setting for simplicity; however, alternative payment models (e.g., prospective payment system), which vary between states, may be used to pay for care provided at CHCs. The costs of HCV antibody (CPT 86803), HCV RNA (qualitative) (CPT 87521), and HCV RNA (quantitative) (CPT 87522) tests reflect the real-world average paid amounts for each CPT code by Medicare, Medicaid, and commercial payers between November 2022 to October 2023 (most recent data available at the time of data extraction). The data were procured from IQVIA’s proprietary healthcare claims remittance database. The cost of a 30-minute outpatient clinic visit (CPT 99203) was derived from the Centers for Medicare & Medicaid Services (CMS) 2024 Physician Fee Schedule.^16^ The Medicare rate was also applied to Medicaid. The reimbursement rate for a 30-minute outpatient clinic visit by commercial payers was derived from a publication by the Congressional Budget Office.^17^ Cost inputs derived from the literature were adjusted to 2024 U.S. dollars using the Medical Care Consumer Price Index.^14^

In the ED setting, HCV diagnostic tests are typically included within a single payment (Ambulatory Payment Classifications (APC)) for the overall ED visit and not reimbursed separately. For the one-year time horizon base-case analysis in the ED setting, the total cost (APC and physician payments) of an ED visit requiring a high level of medical decision making (CPT 99285) was applied; the same rate was assumed across all payer types.^16, 18^ The cost of 12 weeks of oral DAA treatment was applied separately, assuming people diagnosed in the ED would be linked to care outside of the ED. Like the other settings, annual management costs of long-term complications were added for the life-time horizon analysis, assuming care would be provided outside of the ED setting.

Baseline health utilities for PWID without HCV, PWID with HCV, and HCV infected individuals with long-term complications were extracted from literature to calculate the total QALYs gained with each testing strategy.^19^

#### Sensitivity Analysis

One-way sensitivity analyses (OWSA) and probabilistic sensitivity analyses (PSA) were conducted to evaluate the impact of uncertainty on the model results. To investigate the effect of input value variability, we adjusted each variable used in the base-case analysis by ±20% or by the associated 95% confidence intervals reported in the literature, while holding all other variables constant. We plotted the ten ICER results with the widest ranges in a tornado diagram to identify the most influential variables on the percentage of cases successfully treated. PSA was conducted using a Monte Carlo simulation with 1,000 iterations to assess the impact of model and parameter uncertainty on results. Clinical inputs including transition probabilities (transmission model), costs, and utility inputs were assigned log-normal, gamma, and beta distributions, respectively. The simulation was conducted using an ICER willingness-to-pay threshold of $100,000 per QALY gained, which is often referenced in the U.S.^20^ PSA results are presented in the quadrants of cost-effectiveness scatter plots.

#### Scenario Analysis

Scenario analysis was conducted to compare a two-step POC algorithm (two-step POC AB_RNA) against SOC in the CHC, HRC, and MO/SM settings, where an initial POC antibody test is performed, followed by the POC HCV RNA test for persons with detected antibodies.

## Results

### Base-case Analysis Results

In the base-case analysis, the prevalence of active HCV infection was projected to be highest in HRC (25.2%) and MO/SM (24.8%) settings, followed by CHC (16.6%), and ED (16.1%). The one-step POC_RNA testing strategy was projected to identify 93.4% of cases across all settings, while SOC identified fewer cases, at best 68.7% in the HRC setting. The one-step POC_RNA strategy led to increases in: LTC, by 73.4%, 39.5%, 37.6%, and 52.1%; treatment initiation, by 20.1%, 12.1%, 48.9%, and 45.8%; and rates of successful treatment, by 6.3%, 3.6%, 26.4%, and 13.7%, in CHC, ED, HRC, and MO/SM, respectively, when compared to SOC. Table 2 presents detailed results across settings. In addition, one-step POC_RNA lowered the rates of forward transmission by 22.0%, 16.3%, 53.3%, and 33.6% in CHC, ED, HRC, and MO/SM, respectively, when compared to SOC. Analysis of long-term outcomes found that the one-step POC_RNA testing strategy resulted in decreased incidence of chronic hepatitis C, cirrhosis and HCC compared to SOC, along with average gains in 0.6 to 1.1 LYs and 0.5 to 1.0 QALYs across care settings.

**Table 2:** Base-case Analysis Results

| **CHC** | **One-step**<br>**POC_RNA**<br>**Relative %**<br>**(Absolute %)** | **SOC**<br>**Relative %**<br>**(Absolute %)** | **Difference** | **ICER**<br>**(\$ / clinical**<br>**outcome unit)** |
| --- | --- | --- | --- | --- |
| **CHC One-year outcomes** |  |  |  |  |
| HCV cases identified | 93.4% (15.5%) | 37.3% (6.2%) | 56.1% (9.3%) | \$3,875.90 |
| Cases linked to care | 87.8% (14.6%) | 14.4% (2.4%) | 73.4% (12.2%) | \$2,964.98 |
| Cases initiating treatment | 22.9% (3.8%) | 2.9% (0.5%) | 20.1% (3.3%) | \$10,828.35 |
| Cases treated successfully | 7.2% (1.2%) | 0.9% (0.1%) | 6.3% (1.1%) | \$34,650.71 |
| Rate of onward transmission (per 1,000 high risk individuals) | 92.8 | 119.0 | -22.0% (-26.2) | \$13,782.33 |
| Total cost (per person tested) | \$604.42 | \$243.43 | \$360.99 | -- |
| Cost of clinic visits (diagnosis) | \$89.85 | \$167.11 | -\$77.27 | -- |
| Cost of testing | \$46.18 | \$18.63 | \$27.54 | -- |
| Cost of HCV treatment | \$468.40 | \$57.68 | \$410.72 | -- |
| **CHC Long-term outcomes** |  |  |  |  |
| Incidence of chronic HCV (%) | 47.8% (7.9%) | 61.8% (10.3%) | -14.1% (-2.3%) | DOMINANT |
| Incidence of cirrhosis including decompensated cirrhosis (%) | 15.4% (2.6%) | 19.9% (3.3%) | -4.5% (-0.8%) | DOMINANT |
| Incidence of HCC (%) | 2.4% (0.4%) | 3.1% (0.5%) | -0.7% (-0.1%) | DOMINANT |
| Mortality rate (per 1,000 PYs) | 21.2 | 24.3 | -3.0 | DOMINANT |
| Median LYs | 21.3 | 20.7 | 0.6 | DOMINANT |
| Median QALYs | 18.0 | 17.5 | 0.6 | DOMINANT |
| Total cost (per person tested) | \$12,145.66 | \$15,174.86 | -\$3,029.20 | -- |
| **ED** | **One-step**<br>**POC_RNA**<br>**Relative %**<br>**(Absolute %)** | **SOC**<br>**Relative %**<br>**(Absolute %)** | **Difference** | **ICER**<br>**(\$ / clinical**<br>**outcome unit)** |
| **ED One-year outcomes** |  |  |  |  |
| HCV cases identified | 93.4% (15.0%) | 34.8% (5.6%) | 58.6% (9.4%) | DOMINANT |
| Cases linked to care | 48.6% (7.8%) | 9.1% (1.5%) | 39.5% (6.3%) | DOMINANT |
| Cases initiating treatment | 14.1% (2.3%) | 2.0% (0.3%) | 12.1% (1.9%) | DOMINANT |
| Cases treated successfully | 4.2% (0.7%) | 0.6% (0.1%) | 3.6% (0.6%) | DOMINANT |
| Rate of onward transmission (per 1,000 high risk individuals) | 95.1 | 113.6 | -16.3% (-18.5) | DOMINANT |
| Total cost (per person tested) | \$1,063.76 | \$1,499.18 | -\$435.42 | -- |
| Cost of ED visit (diagnosis) | \$785.09 | \$1,460.27 | -\$676.18 | -- |
| Cost of testing | \$0.00<br>(APC bundle) | \$0.00<br>(APC bundle) | \$0.00 | -- |
| Cost of HCV treatment | \$278.67 | \$38.91 | \$239.76 | -- |
| **ED Long-term outcomes** |  |  |  |  |
| Incidence of chronic HCV (%) | 48.8% (7.9%) | 62.5% (10.0%) | -13.6% (-2.2%) | DOMINANT |
| Incidence of cirrhosis including decompensated cirrhosis (%) | 15.4% (2.5%) | 20.1% (3.2%) | -4.4% (-0.7%) | DOMINANT |
| Incidence of HCC (%) | 2.4% (0.4%) | 3.2% (0.5%) | -0.7% (-0.1%) | DOMINANT |
| Mortality rate (per 1,000 PYs) | 21.1 | 24.0 | -2.9 | DOMINANT |
| Median LYs | 21.3 | 20.7 | 0.6 | DOMINANT |
| Median QALYs | 18.0 | 17.5 | 0.5 | DOMINANT |
| Total cost (per person tested) | \$11,888.83 | \$15,264.93 | -\$3,376.10 | -- |
| **HRC** | **One-step**<br>**POC_RNA**<br>Relative %<br>(Absolute %) | **SOC**<br>Relative %<br>(Absolute %) | **Difference** | **ICER**<br>(\$ / clinical<br>outcome unit) |
| **HRC One-year outcomes** |  |  |  |  |
| HCV cases identified | 93.4% (23.5%) | 68.7% (17.3%) | 24.7% (6.2%) | \$27,928.66 |
| Cases linked to care | 87.8% (22.1%) | 50.2% (12.6%) | 37.6% (9.5%) | \$18,307.55 |
| Cases initiating treatment | 86.2% (21.7%) | 37.3% (9.4%) | 48.9% (12.3%) | \$14,085.62 |
| Cases treated successfully | 46.6% (11.7%) | 20.2% (5.1%) | 26.4% (6.7%) | \$26,068.70 |
| Rate of onward transmission (per 1,000 high risk individuals) | 60.0 | 128.5 | -53.3% (-69) | \$25,323.68 |
| Total cost (per person tested) | \$3,262.60 | \$1,527.08 | \$1,735.52 | -- |
| Cost of clinic visits (diagnosis) | \$82.28 | \$153.05 | -\$70.76 | -- |
| Cost of testing | \$34.32 | \$20.33 | \$13.98 | -- |
| Cost of HCV treatment | \$3,146.00 | \$1,353.70 | \$1,792.31 | -- |
| **HRC Long-term outcomes** |  |  |  |  |
| Incidence of chronic HCV (%) | 31.8% (8.0%) | 47.5% (12.0%) | -15.7% (-4.0%) | DOMINANT |
| Incidence of cirrhosis including decompensated cirrhosis (%) | 10.2% (2.6%) | 15.3% (3.9%) | -5.1% (-1.3%) | DOMINANT |
| Incidence of HCC (%) | 1.6% (0.4%) | 2.4% (0.6%) | -0.8% (-0.2%) | DOMINANT |
| Mortality rate (per 1,000 PYs) | 21.5 | 26.6 | -5.1 | DOMINANT |
| Median LYs | 21.2 | 20.2 | 1.1 | DOMINANT |
| Median QALYs | 18.0 | 17.0 | 1.0 | DOMINANT |
| Total cost (per person tested) | \$14,181.18 | \$18,339.51 | -\$4,158.33 | -- |
| **MO/SM** | **One-step RNA**<br>**POC**<br>Relative %<br>(Absolute %) | **SOC**<br>Relative %<br>(Absolute %) | **Difference** | **ICER**<br>(\$ / clinical<br>outcome unit) |
| **MO/SM One-year outcomes** |  |  |  |  |
| HCV cases identified | 93.4% (23.2%) | 61.2% (15.2%) | 32.2% (8.0%) | \$16,812.43 |
| Cases linked to care | 87.8% (21.8%) | 35.7% (8.9%) | 52.1% (12.9%) | \$10,384.20 |
| Cases initiating treatment | 66.2% (16.4%) | 20.4% (5.1%) | 45.8% (11.4%) | \$11,815.43 |
| Cases treated successfully | 19.9% (4.9%) | 6.1% (1.5%) | 13.7% (3.4%) | \$39,384.76 |
| Rate of onward transmission (per 1,000 high risk individuals) | 140.3 | 211.2 | -33.6% (-70.9) | \$18,937.69 |
| Total cost (per person tested) | \$2,152.72 | \$810.06 | \$1,342.65 | -- |
| Cost of clinic visits (diagnosis) | \$91.94 | \$171.00 | -\$79.06 | -- |
| Cost of testing | \$49.43 | \$23.65 | \$25.78 | -- |
| Cost of HCV treatment | \$2,011.35 | \$615.41 | \$1,395.94 | -- |
| **MO/SM Long-term outcomes** |  |  |  |  |
| Incidence of chronic HCV (%) | 43.4% (10.8%) | 55.0% (13.6%) | -11.6% (-2.9%) | DOMINANT |
| Incidence of cirrhosis including decompensated cirrhosis (%) | 14.0% (3.5%) | 17.7% (4.4%) | -3.7% (-0.9%) | DOMINANT |
| Incidence of HCC (%) | 2.2% (0.5%) | 2.8% (0.7%) | -0.6% (-0.1%) | DOMINANT |
| Mortality rate (per 1,000 PYs) | 25.2 | 28.8 | -3.6 | DOMINANT |
| Median LYs | 20.5 | 19.7 | 0.8 | DOMINANT |
| Median QALYs | 17.3 | 16.6 | 0.7 | DOMINANT |
| Total cost (per person tested) | \$18,139.76 | \$21,326.52 | -\$3,186.76 | -- |

CHC=community health centers; ED=emergency departments; HRC=harm reduction centers; MO/SM=mobile outreach / street medicine programs; SOC=standard of care; POC=point-of-care; ICER=incremental cost-effectiveness ratio; HCC=hepatocellular carcinoma; PY=person-years; LY=life-years; QALY=quality-adjusted life-years

From the payer perspective across CHC, HRC, and MO/SM settings, adopting one-step POC_RNA resulted in increased HCV identification and the percentage of cases in each step of the care cascade. The combined benefits of one-step POC_RNA were associated with initial cost increases over a one-year time horizon of $361, $1,736, and $1,343 per person tested in CHC, HRC, and MO/SM settings, respectively, when compared to SOC. The higher upfront costs of the one-step POC_RNA testing strategy were mainly attributed to higher treatment costs due to increased rates of treatment initiation with DAAs. The POC_RNA strategy had smaller increases in testing costs ($30 per person), which was offset by cost-savings observed with fewer clinic visits (approximately -$70 per person across these settings) relative to SOC.

Most EDs currently do not have a HCV test-and-treat infrastructure. The model projected that 12.1% more diagnosed people would be initiated on treatment with the one-step POC_RNA strategy, compared to SOC in EDs. The one-step POC_RNA strategy was projected to yield cost-savings of $435 per person tested within a one-year time horizon in the ED setting. Therefore, POC_RNA is considered a dominant strategy over SOC in the ED setting because it is associated with lower costs and improved testing outcomes.

Over the life-time horizon, one-step POC_RNA testing averted the development of long-term HCV complications and prolonged survival (increased life-years) among the model cohort, compared to SOC. The one-step POC_RNA strategy resulted in lower total costs than the SOC strategy with average cost savings of $3,387 per person tested. Therefore, the POC_RNA testing strategy is the dominant strategy across all settings over a life-time horizon because the short-term test-and-treat benefits reduced the incidence of chronic HCV infections and complications, which resulted in long-term cost-savings to U.S. payers.

### Sensitivity Analyses Results

The tornado diagrams generated in the one-way sensitivity analyses for the CHC (Supplement Figure 1a), HRC (Supplement Figure 1c), and MO/SM (Supplement Figure 1d) settings show that the ICER (cost / % of diagnosed cases successfully treated) was most sensitive to the specificity input of the POC HCV RNA test, with lower test specificity leading to higher incremental costs. In the ED setting (Supplement Figure 1b), the ICER was most sensitive to the percentage of patients initiated on treatment after diagnosis by one-step POC_RNA, the percentage of patients linked to care after diagnosis by one-step POC_RNA, and the overall HCV prevalence in the ED setting. As expected, lower costs are associated with lower input values of each of these three variables due to reduced treatment costs, which appear to strengthen the dominance of the POC_RNA strategy. However, if the objective is to improve HCV test-and-treat outcomes, increases in all three input values would be necessary. The threshold value at which the POC_RNA strategy changes from being cost-savings to being more expensive than SOC in the one-year time horizon is when 75% of cases linked to care are initiated on treatment in the ED setting.

Probabilistic sensitivity analyses (Supplement Figures 2a-d) projected that the one-step POC_RNA strategy would be dominant over SOC in 95%, 96%, 82%, and 86% of the iterations in the CHC, ED, HRC, and MO/SM settings, respectively, at a willingness-to-pay threshold of $100,000 per QALY.

### Scenario Analysis Results

When a two-step POC AB_RNA algorithm was assessed against SOC, the POC AB_RNA algorithm identified fewer cases (−3.8% in CHC, -7.1% in HRC, -6.3% in MO/SM) compared to SOC, due to the lower sensitivity of POC antibody and POC RNA tests compared to laboratory-based tests. As shown in Supplement Table 1, although the two-step POC AB_RNA strategy identified fewer cases, it was projected to increase LTC (7.8% to 15.9%) and treatment initiation (5.4% to 19.6%) across the evaluated settings. These benefits were associated with increased costs per person tested for a two-step POC AB_RNA algorithm of approximately $50 in CHC, $640 in HRC, and $505 in MO/SM, compared to SOC in a one-year time horizon. The ICERs for the long-term outcomes showed mixed results across care settings, with the two-step POC AB_RNA strategy being dominant over SOC across all long-term outcomes in the HRC and MO/SM settings but varying between outcomes in the CHC setting.

## Discussion

Early detection and timely treatment initiation are critical to prevent chronic HCV infection and long-term complications. Our study shows that a one-step POC_RNA testing strategy offers significant benefits over current two-step SOC by enabling immediate HCV status confirmation and faster LTC, particularly in settings with higher HCV RNA prevalence such as CHCs, EDs, HRCs, and MO/SM programs. One-step POC_RNA was projected to increase diagnosis of HCV cases between approximately 25% and 59% relative to SOC, depending on the setting of care. Moreover, increasing the rate of early detection was projected to reduce forward HCV transmission between 22% to 53% among people diagnosed across all settings. Rates of chronic hepatitis C, cirrhosis, and HCC were also reduced, resulting in improvements in the duration and quality of life.

The one-step POC_RNA strategy was also projected to yield longer-term cost-savings to U.S payers and was dominant (lower costs and improved outcomes) over SOC across all care settings over the life-time horizon. Specifically, total costs (per person tested) over the life-time horizon were reduced on average by $3,029 in CHCs, $3,376 in EDs, $4,158 in HRCs, and $3,187 in MO/SM programs. These cost reductions were primarily due to the higher proportion of people successfully diagnosed and treated with one-step POC_RNA, leading to lower incidence of chronic HCV complications and associated medical costs. The cost reductions subsequently offset the higher initial costs associated with POC HCV RNA testing attributed to increased numbers of people initiated on treatment. As a result, one-step POC_RNA emerged as the dominant strategy across all the care settings analyzed, offering substantial clinical and economic benefits over SOC.

Most EDs do not currently focus on HCV diagnosis or have an effective HCV test-and-treat infrastructure. Manteuffel, et al. assessed the impact of implementing a reflex testing protocol in a large, urban ED on the rate of screening completion (two-step SOC algorithm) and LTC.^21^ Their study showed that reflex testing increased the percentage of people with positive antibody tests who received HCV RNA testing from 55% to 84%, but it did not significantly impact LTC (10.6% versus 14.2% [p=0.13] linked to care before versus after reflex testing protocol implementation). Notably, HCV antibody positivity was 11.2% and HCV RNA positivity was 62.6%, supporting the hypothesis that EDs are efficient settings to identify cases. Our study suggests that a one-step POC_RNA strategy could effectively increase HCV diagnosis, LTC, and treatment initiation, warranting efforts to enable a HCV test-and-treat infrastructure in EDs as a critical setting of care that could significantly contribute toward reducing the clinical and economic burden of HCV in the U.S.

Shih, et al. conducted a cost-effectiveness analysis from the perspective of the Australian government, evaluating one-step POC_RNA, two-step POC antibody test followed by POC RNA test for those with detected antibodies (POC AB_RNA), and SOC among people at risk of HCV infection. Results of our study differ from Shih et al., who found that, while one-step POC_RNA was cost-effective compared to SOC, the two-step POC AB_RNA strategy was more cost-effective than one-step POC_RNA when HCV antibody prevalence was less than 74%. In contrast, our study found that a one-step POC_RNA diagnostic strategy was the optimal strategy, followed by two-step POC AB_RNA, even when HCV antibody prevalence was as low as 9.2%. Although both our studies used similar methods and assumptions, differences in findings may be due to a few reasons. First, our model design differed with the inclusion of a transmission model to estimate the number of secondary cases resulting from forward transmission. Second, our model included all steps of the HCV care cascade, capturing outcomes for individuals who were undiagnosed, untreated, or who were treated but did not achieve SVR12. Lastly, our analysis included post-diagnosis and post-treatment initiation costs and outcomes from a one-year and life-time horizons, incorporating costs and savings related to treatment and averted chronic HCV complications.

As with any modeling study, our study is limited to the accuracy of assumptions, model structure, and data inputs. Because our model focuses on costs to healthcare payers, the model cohort includes only people with health insurance. However, we recognize that state and local health departments provide HCV testing and treatment for uninsured populations for which the economic and clinical benefits were not captured in this study. Therefore, the potential societal and public health benefits of reducing the prevalence and incidence of HCV is likely underestimated in this study. While our study found that a one-step POC_RNA testing strategy was optimal (dominant) over the life-time horizon, we recognize that the upfront costs associated with new technology adoption, implementation, and increased treatment costs may not be affordable in settings with limited resources or lack of insurance billing capabilities. Our study found that a two-step POC AB_RNA strategy in which only antibody-positive individuals undergo POC HCV RNA testing may be cost-effective compared to SOC in certain settings but is expected to diagnose fewer cases overall than one-step POC_RNA and may miss diagnosing acute HCV infections.

The United States Viral Hepatitis National Strategic Plan aims to eliminate viral hepatitis in the U.S. by 2030.^22^ Achieving this goal requires the expansion of both HCV testing and treatment; however, several barriers must be addressed to ensure that treatment is available for those with HCV-positive test results. Although a one-step POC_RNA diagnostic strategy can greatly improve rates of case identification and LTC, policy changes are needed to expand access to both testing and treatment, such as removing or reducing prior authorization requirements for DAAs, and prescribing restrictions imposed by payers and state health departments, including re-treatment restrictions for people who have previously undergone treatment. Expanding availability of DAAs outside of specialty pharmacies is necessary to reduce the complexity of test-and-treat, especially for people seeking care in non-traditional care settings. The potential value of POC HCV RNA testing may be even greater if these barriers can be eliminated to enable broad implementation of HCV test-and-treat care pathways. In addition to barrier elimination, the infrastructure needed to support sustainable test-and-treat programs and public health reporting must be funded.

Future studies are needed in the specific settings described in this study, as well as rural settings, correctional facilities and post-carceral settings, to understand the real-world clinical utility of simplified one-step POC_RNA testing and the feasibility of implementing expanded test-and-treat protocols. By focusing on diverse high-burden care settings, researchers can identify and address barriers that may impede the achievement of long-term outcomes as projected in our model and the national HCV elimination goals. Key areas of investigation should include evaluating the effectiveness of POC HCV RNA testing in increasing HCV diagnosis rates, the impact of immediate treatment initiation on patient outcomes and forward transmission, and the logistical challenges of integrating POC test-and-treat protocols into existing practice. Additionally, future research should explore the role of policy changes, such as reducing prior authorization requirements, to enhance access to treatment. Through addressing these barriers, the potential of POC HCV RNA test-and-treat could significantly contribute toward reducing the burden of HCV in the U.S., protect against the risk of viral transmission, and aid in achieving HCV elimination goals.

## Statement of Data Availability

The authors confirm that the data supporting the findings of this study are available within the article. All references have been provided. Values obtained from the IQVIA claims data analysis have been provided directly within the article.

**Supplement Figures 1a-d:**
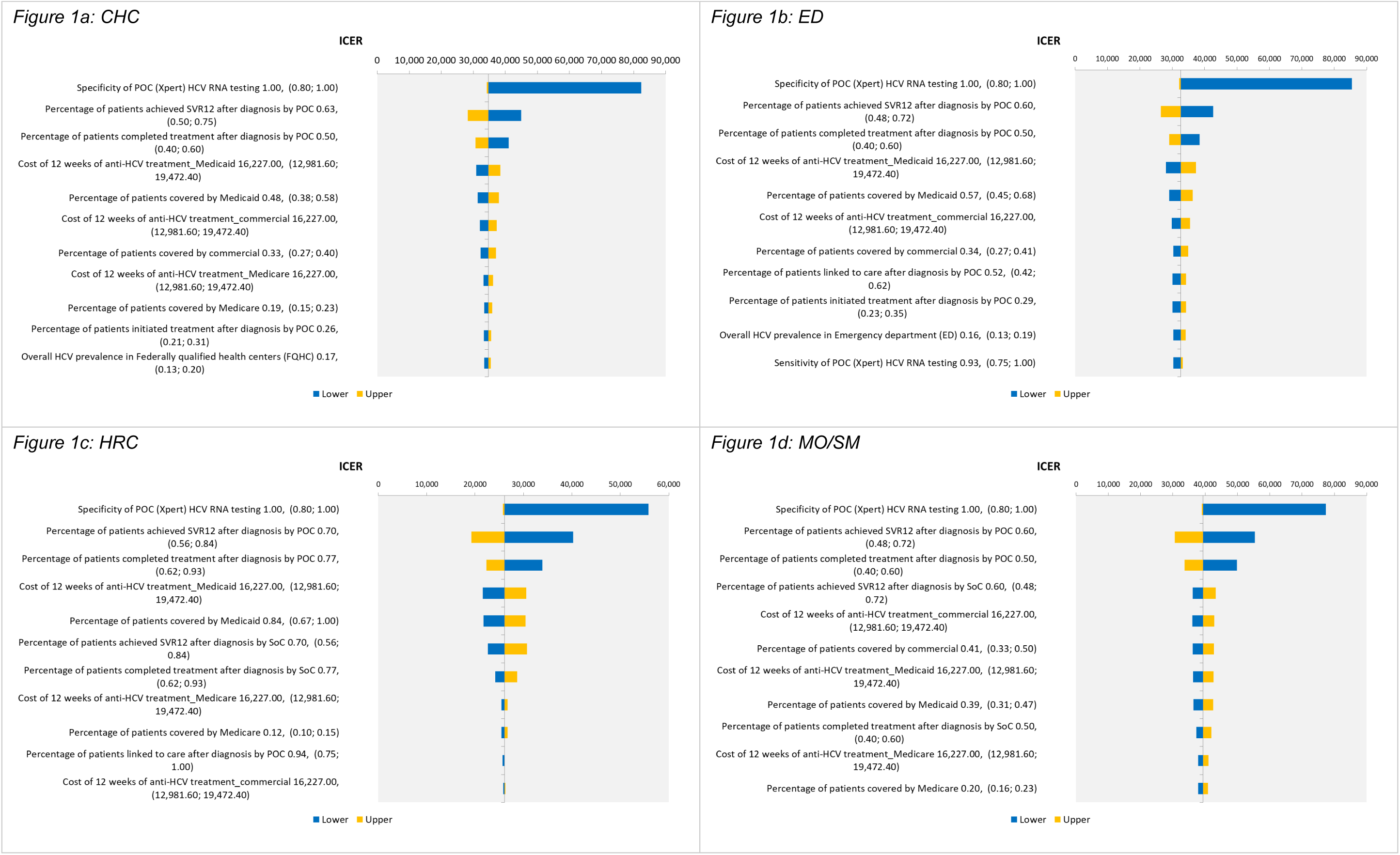
One-way sensitivity analysis by setting; ICER (cost / % treated successfully)

**Supplement Figures 2a-d:**
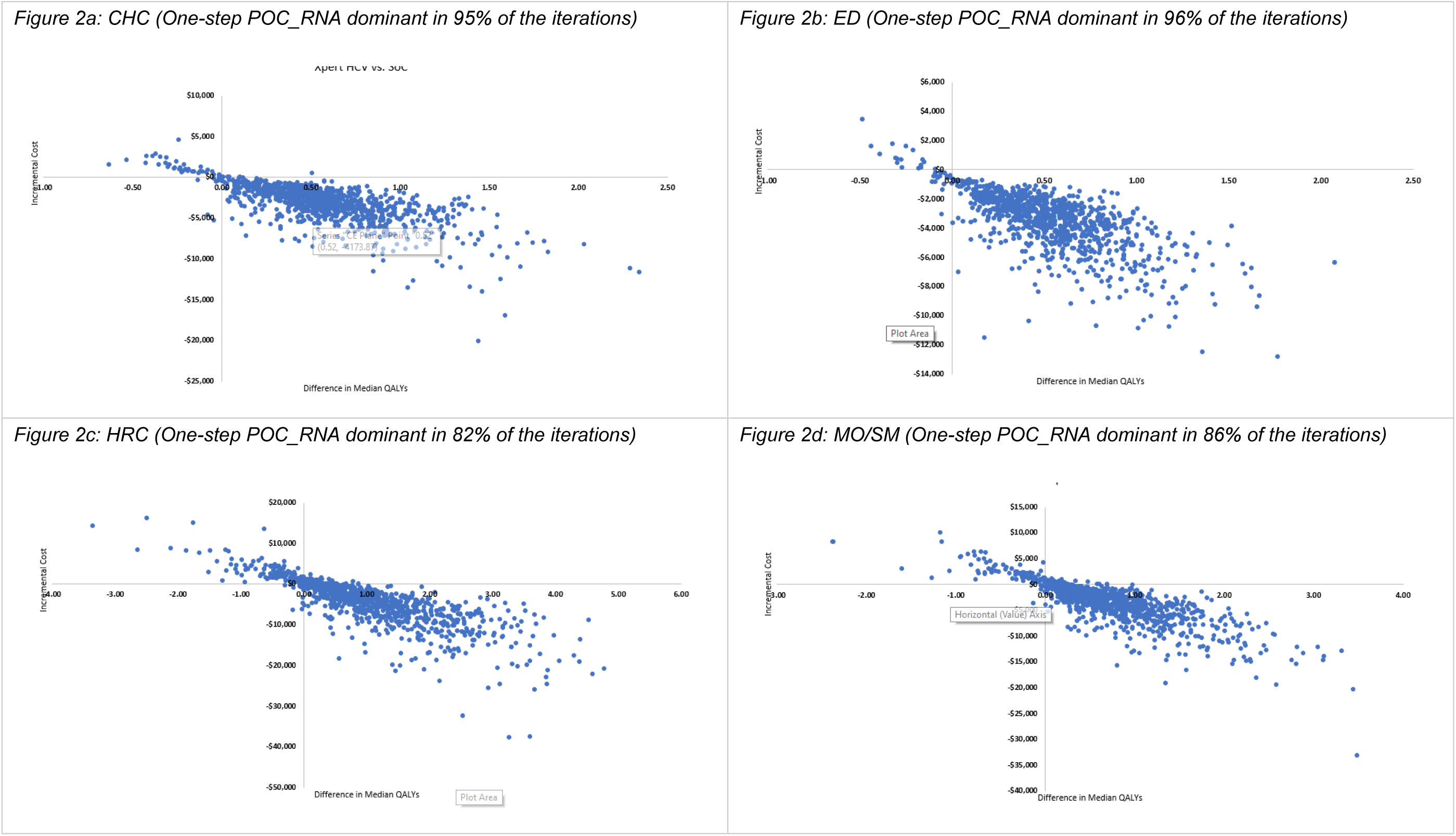
Probabilistic sensitivity analysis by setting (WTP threshold $100,000/QALY)

**Supplement Table 1:** Scenario Analysis Results

| **CHC** | **Two-step POC**<br>**AB_RNA**<br>**Relative %**<br>**(Absolute %)** | **SOC**<br>**Relative %**<br>**(Absolute %)** | **Difference** | **ICER**<br>**(\$ / clinical**<br>**outcome unit)** |
| --- | --- | --- | --- | --- |
| **CHC One-year outcomes** |  |  |  |  |
| HCV cases identified | 33.4% (5.5%) | 37.3% (6.2%) | -3.8% (-0.6%) | Dominated by<br>SOC |
| Cases linked to care | 31.4% (5.2%) | 14.4% (2.4%) | 17.0% (2.8%) | \$1,771.96 |
| Cases initiating treatment | 8.2% (1.4%) | 2.9% (0.5%) | 5.4% (0.9%) | \$5,622.94 |
| Cases treated successfully | 2.6% (0.4%) | 0.9% (0.1%) | 1.7% (0.3%) | \$17,993.40 |
| Rate of onward transmission (per<br>1,000 high risk individuals) | 106.3 | 119.0 | -10.7% (-12.7) | \$3.94 |
| Total cost (per person tested) | \$274.02 | \$224.04 | \$49.98 | -- |
| Cost of clinic visits (diagnosis) | \$89.85 | \$151.07 | -\$61.22 | -- |
| Cost of testing | \$18.30 | \$15.29 | \$3.00 | -- |
| Cost of HCV treatment | \$165.87 | \$57.68 | \$108.20 | -- |
| **CHC Long-term outcomes** |  |  |  |  |
| Incidence of chronic HCV (%) | 62.0% (10.3%) | 61.8% (10.3%) | 0.2% (0.0%) | \$83,780 |
| Incidence of cirrhosis including<br>decompensated cirrhosis (%) | 20.0% (3.3%) | 19.9% (3.3%) | 0.1% (0.0%) | \$259,784 |
| Incidence of HCC (%) | 3.1% (0.5%) | 3.1% (0.5%) | 0.0% (0.0%) | \$1,659,017 |
| Mortality rate (per 1,000 PYs) | 24.3 | 24.3 | 0.1 | \$480 |
| Median LYs | 20.7 | 20.7 | 0.0 | \$2,322 |
| Median QALYs | 17.4 | 17.5 | 0.0 | \$2,775 |
| Total cost (per person tested) | \$15,147.30 | \$15,174.86 | -\$27.56 | -- |
| **ED** | **Two-step POC**<br>**AB_RNA**<br>**Relative %**<br>**(Absolute %)** | **SOC**<br>**Relative %**<br>**(Absolute %)** | **Difference** | **ICER**<br>**(\$ / clinical**<br>**outcome unit)** |
| **ED One-year outcomes** |  |  |  |  |
| HCV cases identified | 54.9% (13.6%) | 61.2% (15.2%) | -6.3% (-1.6%) | Dominated by<br>SOC |
| Cases linked to care | 51.6% (12.8%) | 35.7% (8.9%) | 15.9% (3.9%) | DOMINANT |
| Cases initiating treatment | 38.9% (9.7%) | 20.4% (5.1%) | 18.5% (4.6%) | DOMINANT |
| Cases treated successfully | 11.7% (2.9%) | 6.1% (1.5%) | 5.6% (1.4%) | DOMINANT |
| Rate of onward transmission (per<br>1,000 high risk individuals) | 162.9 | 211.2 | -22.8% (-48.2) | DOMINANT |
| Total cost (per person tested) | \$1,959.80 | \$2,075.68 | -\$115.87 | -- |
| Cost of ED visit (diagnosis) | \$785.09 | \$1,460.27 | -\$675.18 | -- |
| Cost of testing | \$0.00<br>(APC bundle) | \$0.00<br>(APC bundle) | \$0.00 | -- |
| Cost of HCV treatment | \$1,174.71 | \$615.41 | \$559.30 | -- |
| **ED Long-term outcomes** |  |  |  |  |
| Incidence of chronic HCV (%) | 54.4% (13.5%) | 55.0% (13.6%) | -0.6% (-0.2%) | DOMINANT |
| Incidence of cirrhosis including decompensated cirrhosis (%) | 17.5% (4.4%) | 17.7% (4.4%) | -0.2% (0.0%) | DOMINANT |
| Incidence of HCC (%) | 2.7% (0.7%) | 2.8% (0.7%) | 0.0% (0.0%) | DOMINANT |
| Mortality rate (per 1,000 PYs) | 28.7 | 28.8 | -0.1 | DOMINANT |
| Median LYs | 19.8 | 19.7 | 0.0 | DOMINANT |
| Median QALYs | 16.6 | 16.6 | 0.0 | DOMINANT |
| Total cost (per person tested) | \$21,873.70 | \$22,592.14 | -\$718.44 | -- |
| **HRC** | **Two-step POC**<br>**AB_RNA**<br>Relative %<br>(Absolute %) | **SOC**<br>Relative %<br>(Absolute %) | **Difference** | **ICER**<br>(\$ / clinical<br>outcome unit*) |
| **HRC One-year outcomes** |  |  |  |  |
| HCV cases identified | 61.7% (15.5%) | 68.7% (17.3%) | -7.1% (-1.8%) | Dominated by<br>SOC |
| Cases linked to care | 58.0% (14.6%) | 50.2% (12.6%) | 7.8% (2.0%) | \$32,704.75 |
| Cases initiating treatment | 56.9% (14.3%) | 37.3% (9.4%) | 19.6% (4.9%) | \$12,978.92 |
| Cases treated successfully | 30.8% (7.8%) | 20.2% (5.1%) | 10.6% (2.7%) | \$24,020.49 |
| Rate of onward transmission (per 1,000 high risk individuals) | 80.6 | 128.5 | -37.3% (-47.9) | \$13.36 |
| Total cost (per person tested) | \$2,164.80 | \$1,524.37 | \$640.44 | -- |
| Cost of clinic visits (diagnosis) | \$82.28 | \$151.07 | -\$68.79 | -- |
| Cost of testing | \$18.89 | \$19.60 | -\$0.71 | -- |
| Cost of HCV treatment | \$2,063.63 | \$1,353.70 | \$709.93 | -- |
| **HRC Long-term outcomes** |  |  |  |  |
| Incidence of chronic HCV (%) | 44.8% (11.3%) | 47.5% (12.0%) | -2.7% (-0.7%) | DOMINANT |
| Incidence of cirrhosis including decompensated cirrhosis (%) | 14.4% (3.6%) | 15.3% (3.9%) | -0.9% (-0.2%) | DOMINANT |
| Incidence of HCC (%) | 2.3% (0.6%) | 2.4% (0.6%) | -0.1% (0.0%) | DOMINANT |
| Mortality rate (per 1,000 PYs) | 25.8 | 26.6 | -0.9 | DOMINANT |
| Median LYs | 20.4 | 20.2 | 0.2 | DOMINANT |
| Median QALYs | 17.2 | 17.0 | 0.2 | DOMINANT |
| Total cost (per person tested) | \$17,499.90 | \$18,339.51 | -\$839.61 | -- |
| **MO/SM** | **Two-step POC**<br>**AB_RNA**<br>Relative %<br>(Absolute %) | **SOC**<br>Relative %<br>(Absolute %) | **Difference** | **ICER**<br>(\$ / clinical<br>outcome unit*) |
| **MO/SM One-year outcomes** |  |  |  |  |
| HCV cases identified | 54.9% (13.6%) | 61.2% (15.2%) | -6.3% (-1.6%) | Dominated by<br>SOC |
| Cases linked to care | 51.6% (12.8%) | 35.7% (8.9%) | 15.9% (3.9%) | \$12,779.37 |
| Cases initiating treatment | 38.9% (9.7%) | 20.4% (5.1%) | 18.5% (4.6%) | \$10,984.48 |
| Cases treated successfully | 11.7% (2.9%) | 6.1% (1.5%) | 5.6% (1.4%) | \$36,614.94 |
| Rate of onward transmission (per 1,000 high risk individuals) | 162.9 | 211.2 | -22.8% (-48.2) | \$10.47 |
| Total cost (per person tested) | \$1,290.00 | \$785.25 | \$504.75 | -- |
| Cost of clinic visits (diagnosis) | \$91.94 | \$151.07 | -\$59.13 | -- |
| Cost of testing | \$23.35 | \$18.77 | \$4.58 | -- |
| Cost of HCV treatment | \$1,174.71 | \$615.41 | \$559.30 | -- |
| **MO/SM Long-term outcomes** |  |  |  |  |
| Incidence of chronic HCV (%) | 54.4% (13.5%) | 55.0% (13.6%) | -0.6% (-0.2%) | DOMINANT |
| Incidence of cirrhosis including decompensated cirrhosis (%) | 17.5% (4.4%) | 17.7% (4.4%) | -0.2% (0.0%) | DOMINANT |
| Incidence of HCC (%) | 2.7% (0.7%) | 2.8% (0.7%) | 0.0% (0.0%) | DOMINANT |
| Mortality rate (per 1,000 PYs) | 28.7 | 28.8 | -0.1 | DOMINANT |
| Median LYs | 19.8 | 19.7 | 0.0 | DOMINANT |
| Median QALYs | 16.6 | 16.6 | 0.0 | DOMINANT |
| Total cost (per person tested) | \$21,203.89 | \$21,326.52 | -\$122.63 | -- |

CHC=community health centers; ED=emergency departments; HRC=harm reduction centers; MO/SM=mobile outreach / street medicine programs; SOC=standard of care; POC=point-of-care; ICER=incremental cost-effectiveness ratio; HCC=hepatocellular carcinoma; PY=person-years; LY=life-years; QALY=quality-adjusted life-years

## Acknowledgments

The authors would like to thank Dr. Jennifer Rakeman-Cagno, Tamara Ooms, and Dr. Alex Cho of Cepheid, and Dr. Jacob Manteuffel of Henry Ford Hospital for critical review of the manuscript.

## Author Contributions

Neda Al Rawashdh: Model Conceptualization; Methodology; Data Curation; Formal Analysis; Visualization; Writing – original draft

Cheryl P. Ferrufino: Model Conceptualization; Methodology; Data Curation; Formal Analysis; Project Administration; Visualization; Writing – original draft

Mindy M. Cheng: Funding Acquisition; Model Conceptualization; Methodology; Writing – critical review and editing

Shivang Doshi: Model Conceptualization; Writing – critical review and editing

Emily B. Kahn: Model Conceptualization; Methodology; Project administration; Supervision; Writing – critical review and editing

